# Physical calibration and signal periodic updates to synchronize IMUs for 5-day continuous knee flexion tracking

**DOI:** 10.64898/2026.11.14.26368098

**Authors:** Ke Song, Josh R. Baxter

## Abstract

Wearable inertial measurement unit (IMU) sensors provide a promising method to assess real-world human motion kinematics. However, multi-day multi-IMU tracking is difficult due to the desynchronization driven by inaccurate IMU clocks. In this study, 15 volunteers wore IMUs continuously on their thighs and shanks for 5 weekdays (Mon-Fri), which included 3 repeated in-lab motion capture sessions. We synchronized thigh and shank IMUs using two methods. The first was physical calibration, where we maneuvered the IMUs in a rigid holder to produce sync signals pre-donning (Mon) and post-doffing (Fri), then linearly resampled data from one IMU to match the other via cross-correlation. The second was signal-based periodic updates, where we shifted IMU clocks hourly based on the timewise alignment of preprocessed body-worn acceleration signals. After syncing, we used sensor-to-segment alignment and accelerometer-gyroscope fusion to estimate knee flexion and compared results to camera-based motion capture. Physical calibration alone removed clock offset in most cases, where IMU-estimated knee flexion generally matched motion capture (correlation ≥0.944; root-mean-square difference RMSD ≤6.9°). But in some participants, residual mid-week desynchronization (∼0.2s) resulted in misestimated knee flexion (Wed: RMSD up to ∼19°). After applying signal-based updates or combining it with physical calibration, IMU matched motion capture on all 3 days (Mon: r ≥0.975, RMSD ≤6.1°; Wed: r ≥0.971, RMSD ≤6.2°, Fri: r ≥0.977; RMSD ≤5.9°). Our multi-modal synchronization enables week-long knee flexion tracking and lays a foundation for advanced syncing tools, for improving the feasibility of wearable multi-IMU systems to continuously monitor real-world knee kinematics in clinical studies.

## I. Introduction

KNEE mobility is critical for daily function and long-term joint health. Conventional assessments of knee kinematics rely on camera-based motion capture in the lab, which do not capture the frequency and variability of habitual movement in daily life. Wearable inertial measurement units (IMUs) offer a promising alternative by using accelerometer-gyroscope fusion [1], [2] to track segment orientation and joint angles outside the laboratory. Recent studies, including our own [3], have shown that IMUs track knee flexion with accuracy comparable to camera-based motion capture during common movements such as gait [1], [4], [5] and stair navigation [6]. However, nearly all such validation studies were performed in a single experimental session, leaving it unclear whether this tracking quality holds after multiple days of continuous recording – the use case that most multi-day IMU monitoring studies require.

Several studies have recorded multi-day IMU data to report cumulative real-world metrics, most commonly step count and activity level [7], [8], [9] and more recently cumulative kinematics like limb elevation duration or body rotation [10], [11], [12]. A critical technical barrier to extending this work to joint kinematics is inter-sensor synchronization. Unlike single-sensor metrics, multi-IMU measures like the knee flexion angle – an established biomarker in knee osteoarthritis [13], [14] – require that all sensors share a common, accurate clock, which is difficult to guarantee over multiple days [15]. Research-grade high-end IMUs can synchronize in real time via wired triggers or radio-frequency signal checks [16], but these constrain sensor placement, shorten battery life, and increase user burden, making them impractical for large multi-day studies. Lower-cost, unconnected IMUs such as the Axivity AX6 (Axivity Ltd., Newcastle upon Tyne, UK) can log a full week of accelerometer and gyroscope data without recharging, which improves wearer adherence [8], [11] but often sacrifice sensor synchronization features for extended logging capabilities. Such low-cost sensors rely on internal quartz clocks that are inaccurate by tens of parts per million [17] – enough for two AX6 units to desynchronize by several seconds per day [15], far exceeding the precision needed to align thigh and shank motion for knee flexion estimation.

Past studies have addressed this by prescribing wearers to physically maneuver multiple IMUs together to generate a shared calibration signal [11], [12], but participants do not always adhere to these instructions [11]. A simpler strategy is for research staff to perform this calibration maneuver only at the start and end of the wearing period and resample the data assuming a linear inter-IMU clock drift [17]. However, this procedure relies on the linearity of sensor clock drift, which is sensitive to acceleration and temperature [17]. Importantly, calibration-based strategies are easy to implement in theory but challenging in practice. Requiring multiple lab visits or relying on participants to perform the sensor synchronization themselves will result in data attrition. Early sensor stoppage due to battery loss, insufficient memory, and either forgetting or incorrectly performing the maneuver could all disable calibration-based synchronization. Moreover, nearly all prior multi-day kinematic monitoring has targeted the upper limb or torso [10], [11], [12]. To our knowledge, no study has confirmed whether a multi-IMU system can track knee flexion with camera-comparable accuracy across multiple days of continuous wear.

Our goal was therefore to develop simple, reliable post-processing synchronization methods for low-cost IMUs (specifically, the AX6) to continuously quantify the knee flexion angle over multiple days, with accuracy comparable to camera-based motion capture. We expected that, after proper synchronization, IMU-estimated knee flexion would show small magnitude differences and strong waveform agreement with camera-based motion capture across clinically relevant activities throughout the entire tracking duration.

## II. Methods

### A. IMU sensor device

We used Axivity AX6 (Axivity Ltd., Newcastle upon Tyne, UK), which is a low-cost (∼$250 USD), water-resistant research-grade IMU housed in a compact plastic casing with a micro-USB port for charging and data offload (Fig. 1a). At a 100 Hz logging rate with both accelerometer (16g range) and gyroscope (2000 dps range) active, a fully charged AX6 records approximately 7 days of continuous data without wearer interaction, including recharging. We used these settings (16g, 2000 dps, 100 Hz) for both thigh and shank sensors in all experiments. For each participant, we programmed the thigh and shank IMUs to begin recording simultaneously at a specified time on the day of their first lab visit (always a Monday), shortly before sensor calibration and attachment.

**Fig. 1.**
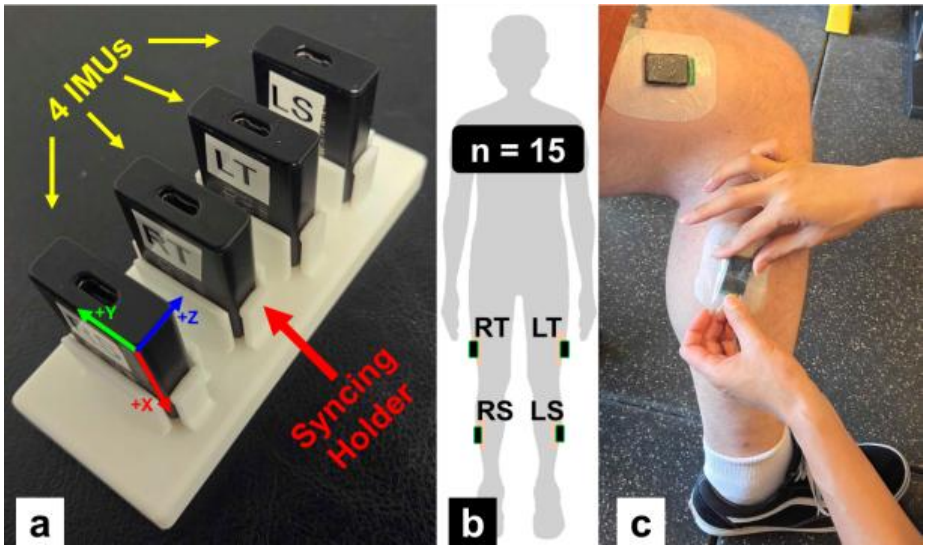
**(a)** 4 IMUs to be worn by a participant, inserted in our custom-made rigid holder. **(b)** IMU attachment locations on the legs. L: Left, R: Right, T: Thigh, S: Shank. We chose an arbitrary leg for analysis; bilateral wearing was for a larger study and feasibility testing on wearer comfort. **(c)** IMU attachments showing all 3 tapes: a protective base layer (white opaque), double-sided adhesive (green tab), and a waterproof cover (transparent, its backing being peeled off). Note the base layer shown is larger than the size we typically used (3×2 in).

### B. Study participant characteristics

We recruited 15 healthy young adult volunteers from our local community (8 male, 7 female, 27.1 ± 5.8 years old, body mass index = 24.7 ± 5.8 kg/m^2^) between September 2025 and March 2026. Participants had no current or self-reported lower-limb or back injury in the prior 6 months that could interfere with knee movement in daily living and were required to be able and willing to visit our lab 3 times in one week (Monday, Wednesday, Friday) for motion capture and to wear IMUs on their thighs and shanks continuously across this 5-day span. All participants provided written informed consent. This study was approved by the University of Pennsylvania Institutional Review Board (Protocol #850424) and performed in accordance with the Declaration of Helsinki.

### C. Experimental procedure: IMU 5-day monitoring and 3 repeated in-lab motion capture sessions

*Sensor attachment*. Each participant’s initial visit occurred on a Monday, a mid-week visit that Wednesday, and a final visit that Friday, at approximately the same time each day. We attached IMUs to both legs on Monday and removed them on Friday. We secured a total of 4 AX6 IMUs to each participant – one pair per leg, on the lateral mid-thigh and lateral mid-shank – using a 3-layer medical-grade adhesive system (protective base layer, double-sided bonding tape, and a waterproof cover layer; full product details and application steps in Supplemental Methods S1). Sensor orientation (charging port down, text facing the skin) was matched across all 4 IMUs, which was necessary for extracting calibration syncing signals (Section E) and sensor-to-segment alignment (Section F). Participants replaced the outer waterproof cover as needed but did not replace the base or bonding layers; they were instead instructed to abort the experiment if those layers detached, which occurred for only 1 of 15 participants (later re-enrolled for a successful 5-day monitoring period). For the remaining 14, all sensors stayed at their original locations for the full week.

#### In-lab motion capture

After sensor attachment, each participant performed a static pose trial (for sensor-to-segment alignment; Section F) followed by 6 movement trials common to daily living – overground straight walking, improvised arbitrary walking, double-legged squat, stair step-up, stair step-down, and 30-second sit-to-stand – at a self-selected pace, recorded with markerless motion capture [18] (specific movement start/end definitions are provided in Supplemental Table S1). We recorded at least 3 gait cycles of straight walking and 3–5 repetitions of each other trial. This protocol was repeated on Wednesday and Friday. After the Friday session, we removed all sensors, performed the end-of-monitoring physical calibration (Section E), and offloaded the 5-day IMU data. Participants then completed an exit survey on wearing experience and a diary of notable high-intensity activity during the week (Results are summarized in Appendix A). Briefly, all 15 participants found the 5-day bilateral wear feasible, and 11/15 (73%) would wear sensors for a full week in future studies.

### D. Preparation of IMU data for post-processing

Our IMU processing pipeline converted thigh and shank accelerometer-gyroscope data into knee flexion angle in 3 steps: inter-sensor synchronization, sensor-to-segment realignment, and accelerometer-gyroscope fusion (Fig. 2). We performed all data processing using Python 3.11.5. Although IMUs were worn bilaterally (for a larger ongoing study and for feasibility assessment; Appendix A), we report findings from one leg per participant: the dominant leg for 13 of 15 participants, and the non-dominant leg for the other 2 participants because of sensor recording errors on the dominant leg.

**Fig. 2.**
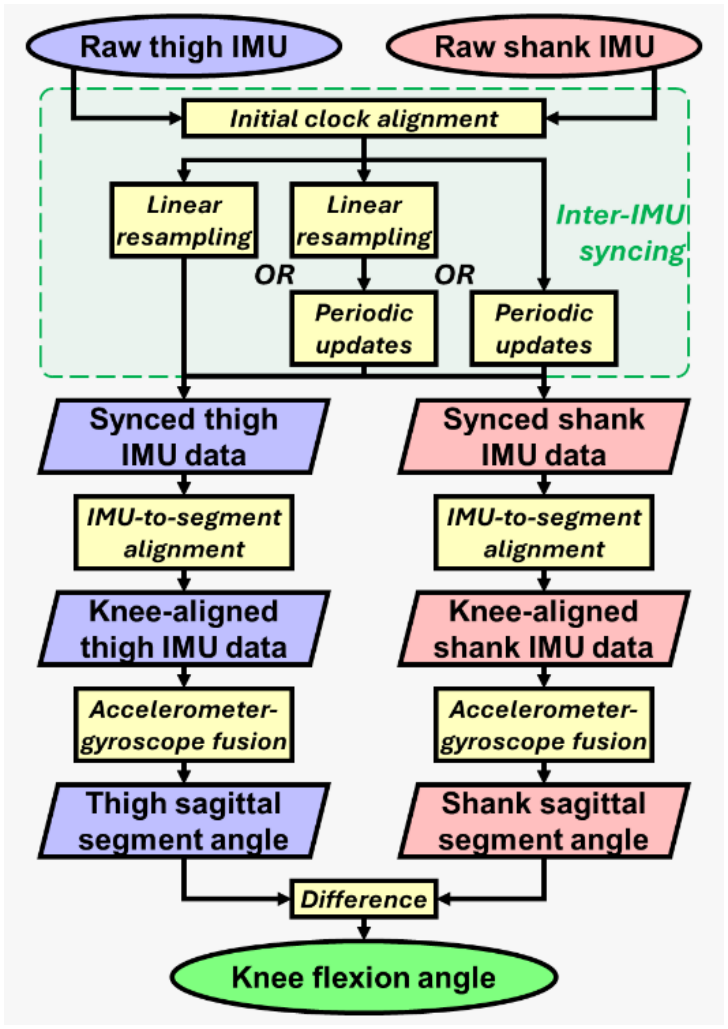
Overview of our workflow to convert thigh and shank IMU data into knee flexion angle. After initial sync alignment, thigh and shank IMUs were synced by either linear resampling (via physical calibration), periodic updates (via body-worn signals), or a combination of both. After syncing, alignment and fusion procedures were performed independently on thigh and shank IMUs. The thigh and shank segment sagittal angles were subtracted at the end to determine the knee flexion angle.

Original time stamps logged by the AX6 were non-uniform.

Therefore, immediately after data import (using the Open Movement Project’s Python module [20]), we resampled each accelerometer and gyroscope channel to an exact 100 Hz using cubic spline interpolation (SciPy). We also removed rare high-magnitude transient error signals (observed once, >100× physical range) that would otherwise interfere with downstream processing that references global signal maxima.

### E. Inter-IMU synchronization: two methods

We developed two independent inter-sensor synchronization methods: (1) calibration-based linear resampling, using syncing signals from a physical calibration tool and maneuver performed before and after the wearing period; and (2) signal-based periodic updates, using engineered features from the body-worn acceleration signals themselves. Both methods began with a shared initial clock alignment, established from the physical calibration signal recorded on Monday. This initial time synchronization does not impose a meaningful burden on experimental setups because it can be performed at the time of IMU software initialization by the researchers rather than study participants immediately before sensor donning.

#### Physical calibration tool and initial alignment

Because AX6 does not synchronize in real time, we designed and 3D-printed a rigid holder to perform initial time synchronization between the 4 IMUs following established recommendations for physical inter-sensor calibration [15], [16]. We inserted all 4 IMUs into the holder facing upward (Fig. 1a) – opposite their on-body orientation – so that a distinctive, automatically detectable signal is produced (Fig. 3). Immediately before the scheduled recording start on Monday, we placed all 4 IMUs into this static “calibration standby” position and left it for at least the first 30 seconds of all recordings, then performed a 3-step rest-swing-rest maneuver (lifting and swinging the holder for ∼10 s, then resting it again) to produce an isolated syncing signal. We repeated this procedure on Friday, immediately after doffing sensors from the participant. We detected each “calibration standby” interval automatically from characteristic accelerometer/gyroscope signatures (full detection criteria in Supplemental Methods S2) and used cross-correlation (SciPy) of the resulting waveforms (Fig. 3) to establish the initial Monday clock alignment, which was used by both synchronization methods below.

**Fig. 3.**
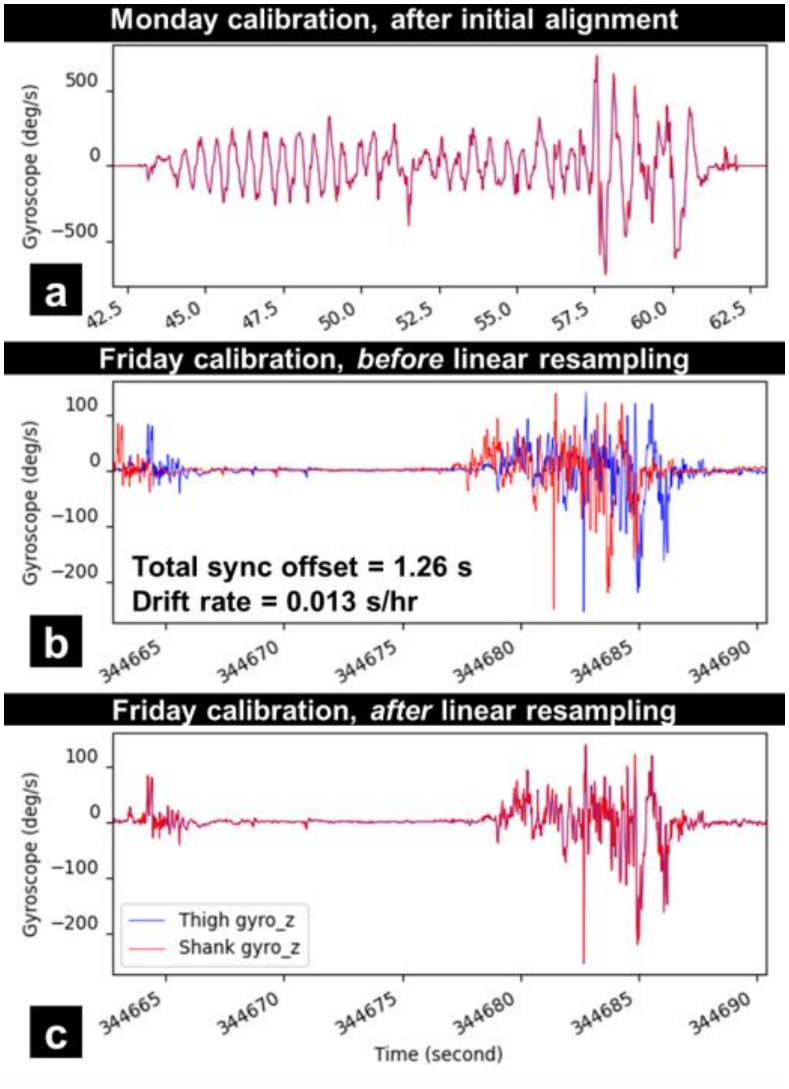
Example of extracted physical calibration sync signals (gyroscope z-axis) after initial alignment and before-after linear resampling. Moving both IMUs while mounted in the rigid-body holder (Fig. 1a) allowed us to capture identical signal waveforms that can be cross-correlated to identify inter-IMU clock offset at the time of the physical calibration. **(a)** In this example, the two IMU clocks were physically calibrated and aligned on Monday, < 1 min after the recordings started. **(b)** At 344,665 seconds (∼4 full days later, on Friday), the two clocks have desynchronized by 1.26 s, or approximately 0.013 s per hour if assuming a linear clock drift. **(c)** Resampling data based on linear dilation of the slower clock (shank IMU in this example, red) resynchronized the two IMUs at the time of the end-of-monitoring calibration performed shortly after Friday’s in-lab session. The linear resampling is only available if both Monday and Friday’s physical calibration signals are recorded by both IMUs. Synchronization quality timewise farther away from initial and final calibration “anchor” timepoints, e.g., on Wednesday, depends on the linearity of the inter-IMU clock drift.

#### Linear resampling via physical calibration

We computed each participant’s average linear inter-IMU clock drift rate from total offset between the Monday and Friday calibration signals (Fig. 3ab) and used it to linearly “dilate” the slower IMU’s time stamps, then resampled all data channels via cubic spline interpolation to a common, uniform 100 Hz clock (Fig. 3c). We identified valid physical calibrations at the experimental beginning and end for all 15 participants. This approach is only available when both the initial and end-of-monitoring calibrations are captured, and its accuracy – particularly mid-week where data is farthest from calibration “anchors” – depends on the assumption that inter-IMU clock drift is linear across the full recording. This was an assumption previously reported to generally hold for AX6 [17], although we did not find it to be universal (Results B & D, Discussion).

#### Periodic updates via body-worn signals

This approach instead assumes that, over sufficiently long windows, thigh and shank acceleration are generally in sync during real-world movement: the shank is moving whenever the thigh is moving, and vice versa. The periodic updates require no physical calibration signal, only concurrent IMU data. We derived a scaled resultant acceleration signal for each sensor as the main syncing signal. We first used a jerk-based algorithm developed in reference to prior work [21] to identify the active daytime periods (as opposed to sleep/rest) (Fig. 4), since acceleration signals during sleep were too quiet to reliably synchronize.

**Fig. 4.**
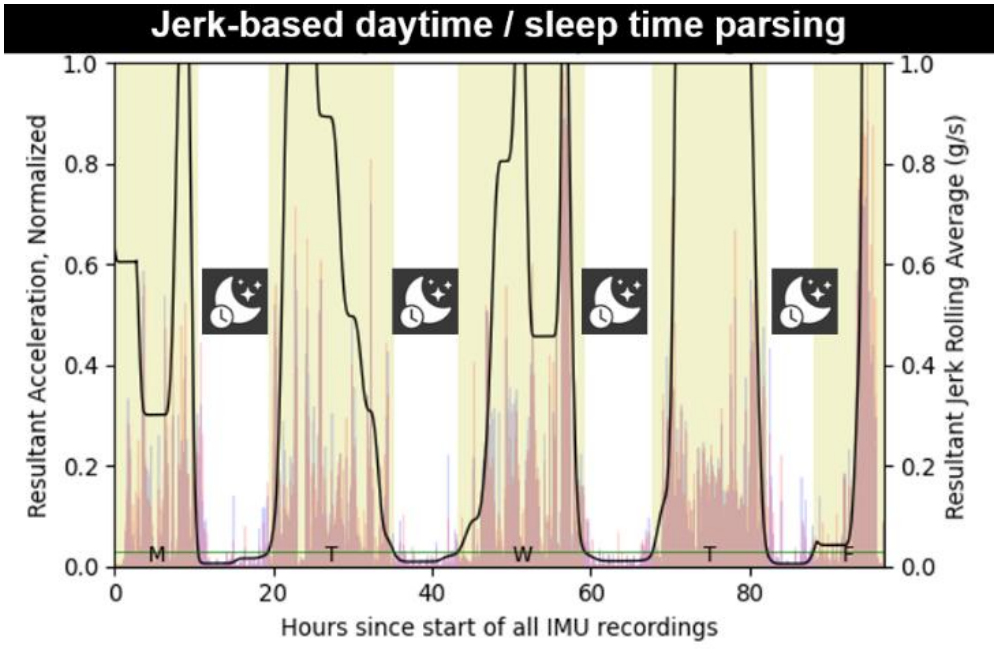
Example of jerk-based daytime and sleep time parsing. Black signal line is the denoised and smoothed sum of jerk absolute value used for thresholding. Thin green horizontal line is the individual-specific threshold. Yellow shaded areas are identified daytime periods (including partially recorded Monday and Friday); unshaded areas in between are sleep periods. Our 1-hour, 4-iteration rolling average filter helped distinguish daytime rests (e.g., mid-Mon, mid-Wed) from true sleep periods. Semitransparent waveforms in the background show normalized resultant accelerations from both IMUs and demonstrate the validity of our daytime parsing algorithm.

Within each daytime hour (Fig. 5), we selected the first sufficiently active 5-minute window, then low-pass filtered (4 Hz cutoff) and demeaned the thigh and shank signals within that window to emphasize their shared, general motion envelope over sensor-specific waveform features (Fig. 6). We then cross-correlated them to estimate that hour’s clock offset, discarding implausibly large offsets (full parameterization in Supplemental Methods S2). We shifted the faster IMU’s data to match the slower one and iterated this check hourly across the full 5-day recording (Fig. 5).

**Fig. 5.**
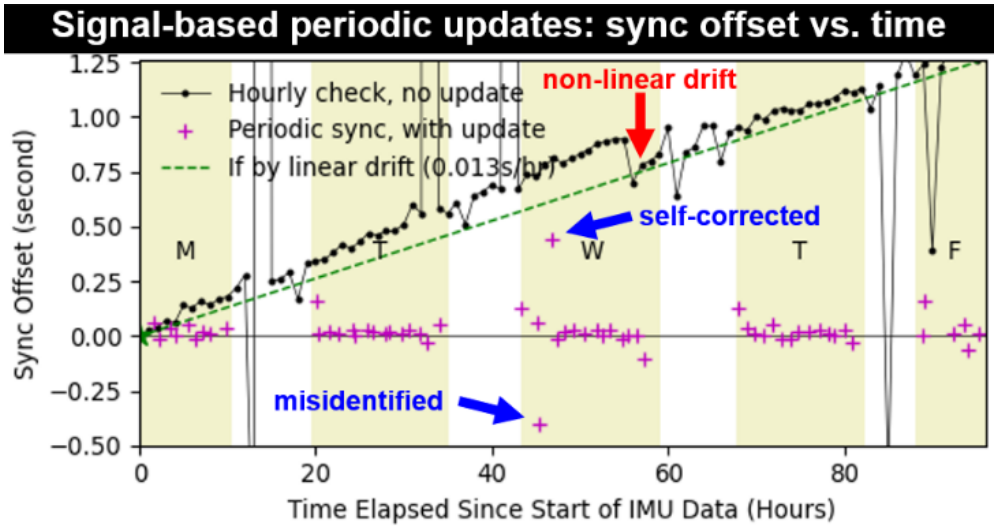
Example from one participant on hourly clock offsets between the thigh and shank IMUs, iteratively identified by the hourly clock updates (“+” crosshairs). The updates, which were larger at the first checkpoint of each day as we expected, kept desynchronization <0.05 s for most of the time. Although misidentifications occasionally occurred, they were usually self-corrected at the next hourly checkpoint (blue arrows). For context, the black dots illustrate offsets of the same IMU pair identified with no day-night parsing (2), criteria-based bout selection (4), filtering (5), or correction. A notable non-linear drift occurred (red arrow), which led to residual desynchronization on much of Wednesday when only using linear resampling for syncing (green dashed line = assumed linear drift correction). There were also substantially more instances of misidentified sync offsets, especially during nighttime (unshaded areas).

**Fig. 6.**
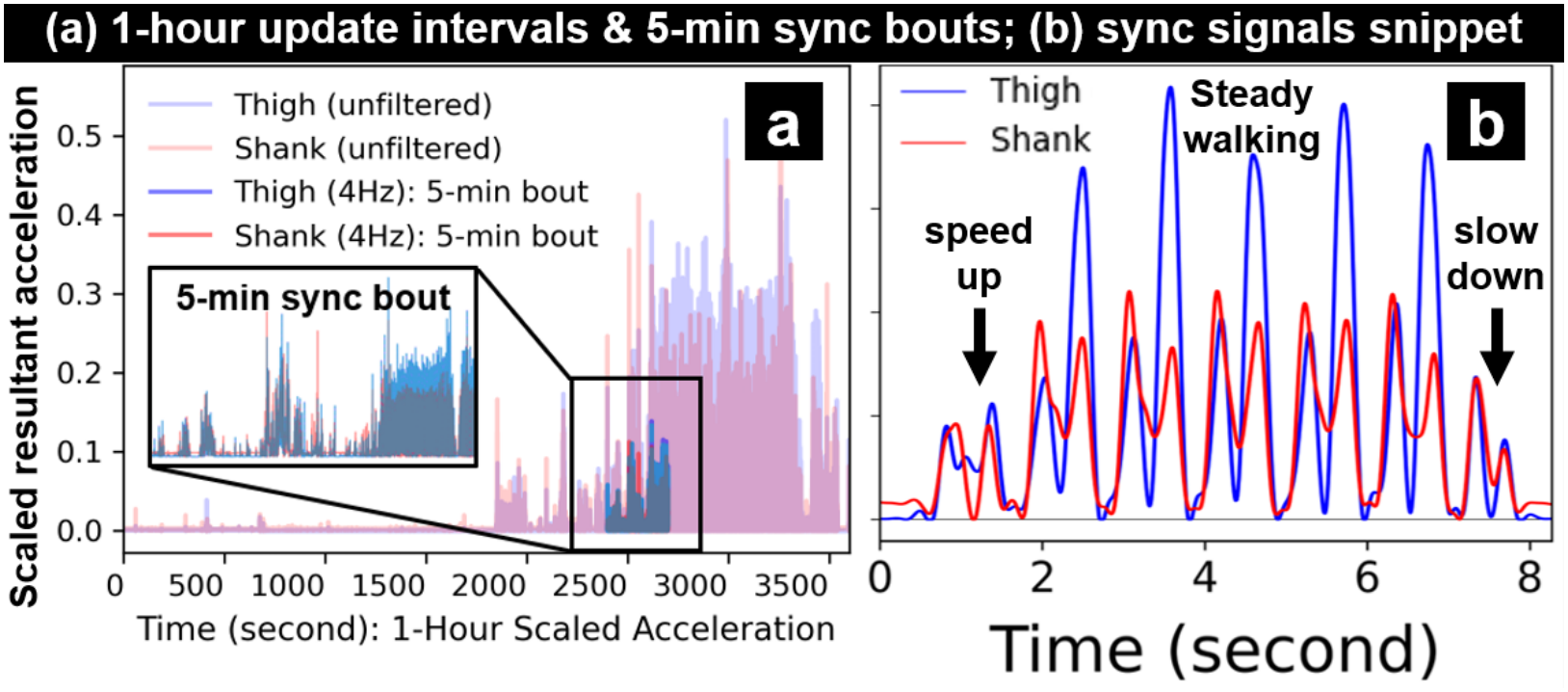
**(a)** Example of a 1-hour update interval, and its criteria-extracted 5-minute sync bout. Note that signals in the first half of this 1-hour interval were too quiet for cross-correlation. Only the signals in the 5-minute bout were filtered, for syncing purposes only. **(b)** Snippet of scaled and 4 Hz low-pass filtered resultant acceleration signals from shank (red) and thigh IMUs (blue) *after* the clock-shifting synchronization. While shank and thigh accelerations exhibit distinct waveforms, low-pass filtering attenuated their unique waveform features (indicated by the lower magnitudes after filtering) while preserving their shared *general* intensity profiles like the speed-up, steady, and slow-down phases of an isolated walking bout. Our rationale for using the thigh-to-shank cross-correlation for synchronization was based on such shared *general* motion intensity envelopes between the shank and thigh.

#### Combining both methods and post-hoc verification

We additionally tested applying periodic updates to data already synchronized via linear resampling and expected this to further reduce any residual non-linear drift. We evaluated the syncing outcomes for all three workflows (linear resampling alone, periodic updates alone, and combined) by rerunning the periodic-update algorithm on each synchronized dataset to quantify the residual inter-IMU clock offset – ideally near 0 – on IMU data over the timespan of each in-lab session (Results B). Residual offset was defined as any remaining clock offset between IMUs after data had passed through synchronization.

### F. Knee flexion angle from synchronized thigh and shank IMUs

We performed sensor-to-segment alignment and accelerometer-gyroscope fusion independently on the synchronized thigh and shank data, following our previously validated single-session procedure [3]. Briefly, we virtually rotated each IMU’s local-frame data into an anatomically meaningful frame aligned with the knee flexion axis and gravity. We approximated the sagittal knee flexion axis via principal component analysis of gyroscope data during Monday’s squatting trial (in place of literature-recommended walking trial [22] due to our short in-lab walkway) and the proximal-distal axis from the static standing pose. To correctly extract IMU data during the time periods of the pose and squat trials, we corrected for IMU-to-motion-capture clock offset (independent of the inter-IMU synchronization) by cross-correlating segment sagittal velocity during the squat trial (full axis derivation details in Supplemental Methods S3).

We then estimated each IMU’s orientation using an open-source, accelerometer-gyroscope fusion algorithm (Madgwick filter, no-magnetometer version, β = 0.5) [2], [3]. We applied data fusion across the full 5-day continuous recording rather than only the brief windows concurrent with reference data, as in most prior work [3], [22]. We converted the resulting orientation quaternions to Euler angles and subtracted the sagittal shank and thigh angles to obtain knee flexion angle, sign-corrected per our established convention [3], [22].

Results yielded continuous ∼90–100 hours of knee flexion time series per participant (30–40 million samples).

### G. Comparison between IMU and in-lab motion capture

We matched IMU-estimated knee flexion to corresponding motion capture trial during each lab session using ground-truth camera time stamps. We first removed the residual absolute IMU-to-lab clock offset (independent of the inter-IMU synchronization) via cross-correlation on each day’s 30-second improvised walk trial. We then applied that offset to all trials recorded that day (same-day trials were recorded <15 min apart, so additional drift was assumed negligible).

We then computed, for each trial repetition, the Pearson correlation coefficient (R_xy_) and root-mean-square difference (RMSD) between IMU-estimated and camera-based knee flexion waveforms, averaged across repetitions and then across participants for each lab day. Following Schober et al. [23], we defined R_xy_ ≥ 0.7 as strong and ≥ 0.9 as very strong agreement; consistent with prior literature [24], [25] and our own work [3], [19], we defined RMSD ≤ 5° as a minimal magnitude difference.

### H. Sensitivity analysis: effect of induced desynchronization on estimated knee flexion magnitude

We performed an ancillary sensitivity analysis to isolate the effect of desynchronization on knee flexion angle estimation independent of other errors known to impact joint kinematic estimates like skin artifact, sensor attachment, and algorithm noise [15], [16]. Using each participant’s post-synchronization, post-alignment, pre-fusion data from Monday’s straight walking trial, we induced artificial clock shifts to the shank data (0.01–0.05 s, in 0.01 s increments) while holding the thigh data fixed, re-ran data fusion, and computed the RMSD between the resulting and the original (synchronized) knee flexion estimates. We used this sensitivity analysis to establish what level of synchronization precision is needed to achieve acceptable knee flexion estimates, under the assumption that all errors are explained by temporal offsets. We considered an error of less than 5° an acceptability threshold [3], [25], but this error tolerance should be reconsidered by investigators based on their own research questions and study designs.

## III. Results

### A. Inter-IMU clock drift behaviors

Inter-IMU clock drift was both device- and experiment-specific; the same IMU pair exhibited different drift patterns in multiple experiments. Using our Monday and Friday calibration anchors as the start and end timepoints, total monitoring durations averaged 95.6 ± 2.1 h (range 92.0–99.2), consistent with our intended 4 full days (96 h). Total inter-IMU drift varied widely across participants (0.47–6.95 s; mean 2.76 ± 2.34 s), corresponding to an assumed-linear drift rate of 0.005–0.074 s/h (mean 0.029 ± 0.025 s/h). The largest rate observed (0.074 s/h, ≈20.5 ppm) fell within previously reported ranges for the AX6 [15], [17]. There was no consistent trend in which IMU’s clock ran faster (shank faster in 9/15 participants, thigh faster in 6/15).

### B. Verification of post-synchronization residual clock offsets, according to the periodic update algorithm

Both methods effectively removed inter-IMU clock offset. Across the 45 sessions (15 participants × 3 lab days), periodic updates alone kept residual offset ≤0.03 s in 34 (76%) and ≤0.05 s in 40 (89%) sessions (worst case: 0.11 s, one Friday session). Linear resampling alone kept residual offset ≤0.03 s in 40 (89%) and ≤0.05 s in 41 (91%) sessions but produced the two largest residual offsets of any verification – 0.19 s and 0.18 s, both on Wednesdays. The other sessions >0.05 s were also on Wednesdays for linear resampling alone. Combining linear resampling with periodic updates performed best: 39 (87%) sessions ≤0.03 s and 44 (98%) ≤0.05 s, with only one session (0.08 s, a Wednesday) exceeding 0.05 s.

### C. Sensitivity of estimated knee flexion angle to induced clock desynchronization

IMU-estimated knee flexion diverged progressively from reference result (synchronized) as induced desynchronization increased (Fig. 7). Averaged across participants, RMSD rose from 1.7° at ±0.01 s to 3.4° (±0.02 s), 5.0° (±0.03 s), 6.6° (±0.04 s), and 8.2° (±0.05 s). At ±0.03 s, RMSD ranged 4.0°– 5.6° across participants, and at ±0.05 s, 6.5°–9.2°. Given our 5° acceptability threshold [25], this places the tolerable inter-IMU sync offset at approximately ±0.03 s.

**Fig. 7.**
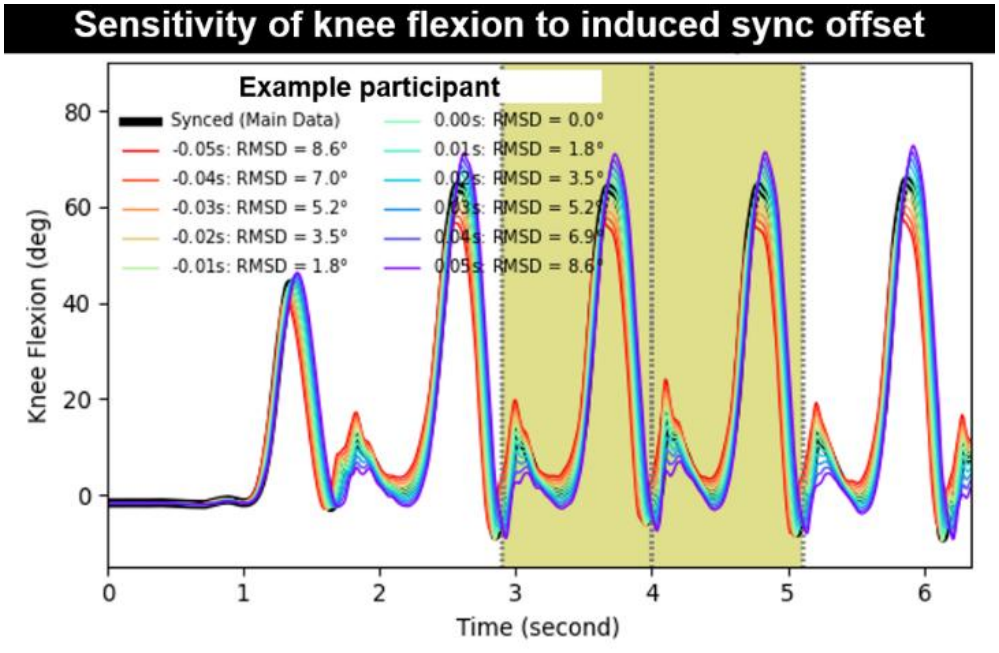
Example on the effect of incrementally induced desynchronization on the estimated knee flexion angle, shown on a straight walking trial from one participant. A ±0.05 s sync offset between the thigh and shank IMUs typically resulted in an 8° – 9° knee flexion difference compared to synced data. To limit potential kinematic inaccuracies under 5°, the inter-IMU sync offset should be controlled within ±0.03 s.

### D. IMU-estimated knee flexion angle vs. camera-based motion capture on all 3 days with in-lab session

Across all 3 in-lab sessions and all three synchronization workflows, IMU-estimated knee flexion showed very strong group-level waveform agreement with camera-based motion capture, with magnitude differences that met or narrowly exceeded our a-priori 5° threshold (full results by syncing workflow, movement type, and day are reported in Table I).

**TABLE I.** Knee Flexion Angle by IMUs vs. Motion Capture: Pearson Correlation & Root-Mean-Square Difference (N = 15)

|  | Pearson Correlation (R<sub>xy</sub>) |  |  | Root-Mean-Square Difference (RMSD) |  |  |
|---|---|---|---|---|---|---|
| Linear Resampling Only |  |  |  |  |  |  |
|  | Monday | Wednesday | Friday | Monday | Wednesday | Friday |
| Straight walking | .975 ± .008 | .944 ± .104 | .977 ± .009 | 6.1° ± 1.4° | 6.3° ± 3.5° | 5.3° ± 1.8° |
| Squat | .996 ± .011 | .984 ± .034 | .997 ± .005 | 5.6° ± 2.9° | 6.4° ± 4.6° | 4.4° ± 1.8° <sup>b</sup> |
| Step up (n = 12)<sup>a</sup> | .994 ± .008 | .982 ± .046 | .995 ± .004 | 5.2° ± 2.4° | 5.3° ± 3.5° | 4.4° ± 2.5° <sup>b</sup> |
| Step down (n = 12)<sup>a</sup> | .994 ± .006 | .985 ± .040 | .996 ± .002 | 6.0° ± 2.3° | 5.4° ± 3.1° | 4.9° ± 2.1° <sup>b</sup> |
| Sit-to-stand | .997 ± .004 | .979 ± .045 | .996 ± .006 | 6.0° ± 3.0° | 6.9° ± 4.8° | 5.1° ± 2.3° |
| Periodic Updates Only |  |  |  |  |  |  |
|  | Monday | Wednesday | Friday | Monday | Wednesday | Friday |
| Straight walking | .979 ± .008 | .971 ± .022 | .977 ± .010 | 5.9° ± 1.5° | 6.2° ± 2.0° | 5.8° ± 2.0° |
| Squat | .996 ± .012 | .995 ± .005 | .994 ± .006 | 5.6° ± 3.0° | 5.3° ± 1.6° | 5.2° ± 1.9° |
| Step up (n = 12)<sup>a</sup> | .995 ± .009 | .993 ± .006 | .991 ± .006 | 4.9° ± 2.6° <sup>b</sup> | 4.9° ± 1.7° <sup>b</sup> | 5.3° ± 2.2° |
| Step down (n = 12)<sup>a</sup> | .995 ± .006 | .995 ± .005 | .996 ± .002 | 5.8° ± 2.4° | 4.9° ± 1.7° <sup>b</sup> | 5.0° ± 2.2° <sup>b</sup> |
| Sit-to-stand | .997 ± .005 | .994 ± .006 | .992 ± .007 | 5.9° ± 3.0° | 5.8° ± 2.3° | 5.9° ± 2.1° |
| Linear Resampling Followed by Periodic Updates |  |  |  |  |  |  |
|  | Monday | Wednesday | Friday | Monday | Wednesday | Friday |
| Straight walking | .975 ± .008 | .977 ± .017 | .978 ± .009 | 6.1° ± 1.5° | 5.5° ± 1.5° | 5.6° ± 1.9° |
| Squat | .995 ± .013 | .996 ± .005 | .996 ± .005 | 5.7° ± 3.0° | 5.0° ± 1.6° <sup>b</sup> | 4.8° ± 1.7° <sup>b</sup> |
| Step up (n = 12)<sup>a</sup> | .994 ± .010 | .994 ± .007 | .992 ± .007 | 5.1° ± 2.5° | 4.6° ± 1.8° <sup>b</sup> | 5.0° ± 2.5° <sup>b</sup> |
| Step down (n = 12)<sup>a</sup> | .994 ± .007 | .996 ± .004 | .995 ± .003 | 6.0° ± 2.3° | 4.7° ± 1.3° <sup>b</sup> | 5.3° ± 2.1° |
| Sit-to-stand | .997 ± .005 | .996 ± .004 | .994 ± .007 | 5.9° ± 2.9° | 5.3° ± 2.1° | 5.4° ± 2.1° |

<sup>a:</sup> Step up and down were analyzed on a subset of 12 participants who completed these trials according to our exact trial definitions.
<sup>b:</sup> Met our a-priori criteria for minimal RMSD between the two systems. All Pearson correlations met the “very strong" criteria.

Linear resampling alone produced very strong agreement on each lab day (R_xy_ ≥ 0.944, RMSD ≤ 6.9°), but this group-level result masked two participants with markedly worse Wednesday tracking (R_xy_: 0.609–0.919, RMSD: 7.0°–18.9°; Fig. 8a) – the only instance where R_xy_ fell below our “strong” criterion. These two outlier sessions were the same two sessions with the largest residual clock offsets identified in our post-hoc verification (Results B) and were the primary source of the elevated group-level variability seen on Wednesday for this method (Table I).

**Fig. 8.**
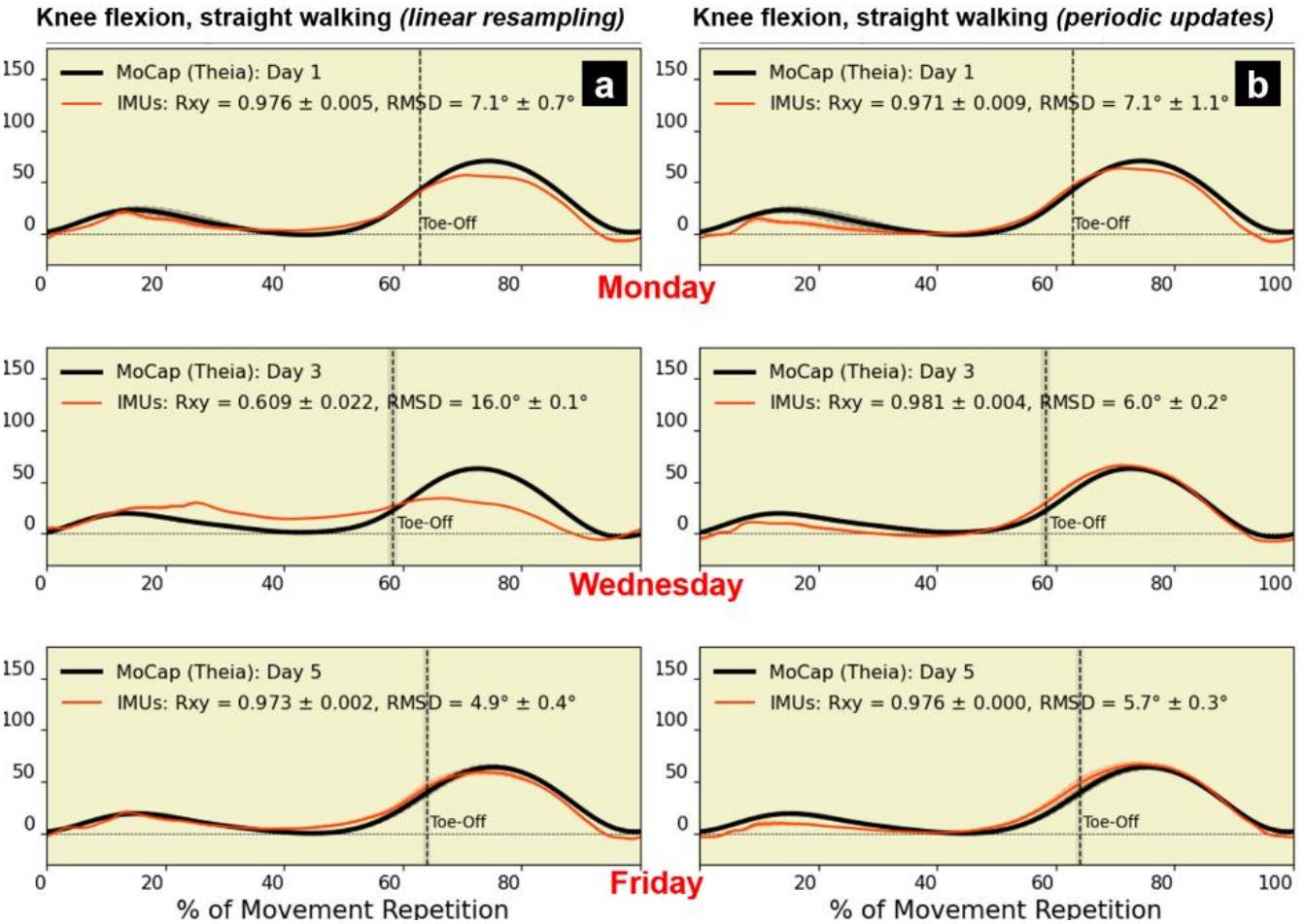
Example knee flexion angle during straight walking gait cycles from a participant (same one as Fig. 5), comparing IMU-based (red) to camera-based estimates (black) on Monday (top), Wednesday (center), and Friday (bottom). (a) Results from data synced by linear resampling *only*. IMUs had reasonable accuracy on Monday (RMSD = 7.1°) and achieved minimal difference to camera-based on Friday (RMSD = 4.9°), but agreements were much worse on Wednesday (R_xy_ = 0.609, RMSD = 16.0°). This is likely explained by the large residual clock offset (0.19s via our verification) caused by non-linear clock drift and unresolved by linear resampling (Fig. 5). (b) Results from data synced by periodic updates *only*. Results greatly improved on Wednesday (RMSD = 6.0°) and remained the same on Monday (RMSD = 7.1°). The marginally worse result on Friday (RMSD = 5.7°) may be due to imperfections in auto-selected sync bouts (Fig. 6a), or that the time of this in-lab trial was farther off its last hourly checkpoint (periodic update) than Friday’s physical calibration (just minutes after the lab session), which allowed minor desynchronization.

Periodic updates alone produced comparable group-level agreement (R_xy_ ≥ 0.971, RMSD ≤ 6.2° across days) but with substantially reduced Wednesday variability, because both linear-resampling outlier sessions tracked well under this method (Fig. 8b); post-hoc verification confirmed their residual offsets fell from 0.19 s and 0.18 s to 0.01 s and 0 s, respectively. Combining both methods produced the most consistent results overall (R_xy_ ≥ 0.975, RMSD ≤ 6.1° across days), with the best Wednesday agreement of the three workflows and no outlier sessions (Table I, Fig. 9).

**Fig. 9.**
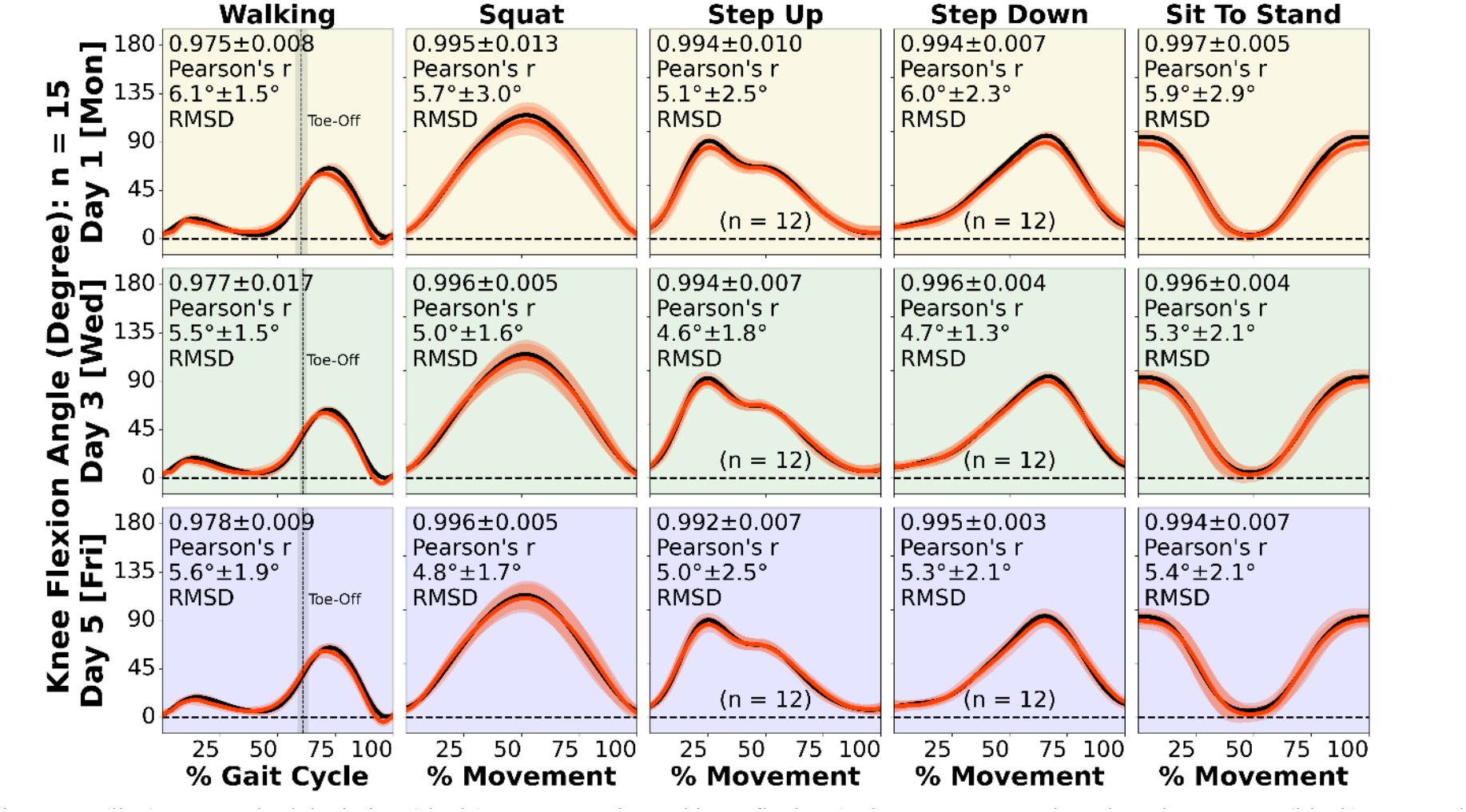
Group-wise mean (line) ± 1 standard deviation (shade) on IMU-estimated knee flexion (red) versus camera-based motion capture (black) on Monday (top), Wednesday (center), and Friday (bottom). Data shown was synced using both calibration-based linear resampling and signal-based periodic updates. Syncing with *either* linear resampling *or* periodic updates produced results of similar agreement to the camera-based reference (Supplemental Material: Fig. S1, S2). However, *only* using linear resampling led to multiple outlier cases where mid-week tracking errors were substantially worse and unacceptable, likely caused by unresolved nonlinear clock drifts (Fig. 8a). In contrast, data synced by periodic updates did not exhibit such failure (Fig. 8b).

## IV. Discussion

Our goal was to develop simple, reliable post-processing synchronization methods that support continuous monitoring of knee flexion over multiple days using low-cost, unconnected IMUs. We developed and compared three workflows – physical calibration-based linear resampling, signal-based periodic updates, and a combination of these strategies – on thigh-shank AX6 pairs worn by 15 healthy adults for 5 days. At the group level, all three workflows produced knee flexion estimates of acceptable accuracy through our previously established kinematic processing pipeline [3]. At the individual level, however, they differed: linear resampling alone produced multiple mid-week outliers, which we traced to non-linear inter-IMU drift that this method cannot correct. Periodic updates fixed this non-linear drift, and combining the two methods further reduced misidentified or unresolved offsets. Knee flexion angle results showed strong agreements with concurrent camera-based motion capture regardless of whether periodic updates were applied on linear resampled or the original IMU data. Our findings therefore generally favor periodic updates over linear resampling, though each method carries its own assumptions and constraints. More broadly, this 5-day validation extends the tracking quality previously demonstrated only in minute-long sessions to ∼100 hours of continuous data, supporting the technical and practical feasibility of multi-day, multi-IMU knee kinematic monitoring in future clinical studies on chronic conditions like knee osteoarthritis.

Consistent with a prior report for the AX6 [17], the inter-IMU drift rates we observed fell within expected ranges.

However, unlike that report, drift was not always linear in our data (13/15 participants, 87%) and varied by experiment even for the same IMU pair, suggesting AX6 clock behavior is not fully device intrinsic. Although device age and ambient temperature are both known to affect quartz-oscillator timing accuracy [26], they were similar across our experiments and sensors (relatively new, <4-year-old). Rather, the two non-linear drift events we observed each coincided with a period of unusually vigorous activity reported in the participant’s activity diaries (a basketball game and an outdoor run). This indicates high-acceleration events, not time or temperature, may be the more relevant driving factor of non-linearity. This observation raises an important caveat for the linear-resampling approach: because it assumes constant drift, it may be less reliable for physically active wearers, even as it performed well for other non-outlier participants and on Monday/Friday even for the two outliers (because calibration “anchors” bound the Monday and Friday sessions directly).

Linear resampling may still be preferable in some contexts because it requires no shared motion between adjacent body segments, thus it remains available for bilateral or otherwise unconnected segments. It may also suit studies with shorter monitoring windows or where sensors can be attached, detached, and recalibrated mid-experiment, since the rigid-holder signal is inherently more reliable than cross-correlating each segment’s naturally distinct motion. Linear resampling may be better suited to less physically active populations, such as many patients with knee osteoarthritis, though this requires direct validation.

Periodic updates, by contrast, kept mid-week data synchronized even in the presence of non-linear drift and produced no poor-tracking outliers at the individual level (Fig. 8). When combined with linear resampling, 98% of sessions stayed within 0.05 s of synchronization. Because our periodic update method uses unlabeled body-worn signals rather than prescribed physical action, it is robust and usable even when an end-of-monitoring calibration is missing or invalid. Also, since it updates forward through the recording, it can still synchronize data up to the point where a sensor stops recording prematurely, a failure mode that would otherwise disable linear resampling entirely. To our knowledge, this is the first method to synchronize thigh and shank IMUs via their shared general motion-intensity envelope. It suggests that valid multi-day knee flexion tracking could in principle extend as long as the paired recordings last, supporting the broader feasibility of week-long multi-IMU kinematic monitoring with low-cost, unconnected sensors.

Our walking RMSD (5°–6°) was modestly larger than the <4° typically reported between IMU and marker-based motion capture [1], [4], [5], which may be partly attributed to our use of markerless reference data (instead of marker-based), though we previously found the two reference systems to closely agree on knee flexion during gait (RMSD <4°) [19]. Camera-based motion capture is itself imperfect relative to ground-truth trackers like biplane fluoroscopy [24]. In one recent study that directly compared IMU-based knee kinematics to fluoroscopy, IMU and marker-based motion capture both showed similar accuracy (RMSD ≈4°) [27]. Combined with our sensitivity analysis that places the 5° error threshold [25] at an approximately 0.03 s sync offset, our group-level RMSD of 5°–6° on all 3 days suggests our synchronization kept the IMU pair within roughly 0.03 s of true alignment throughout the week. This means our synchronization methods were able to extend prior single-session validation – typically limited to minutes of data – to continuous tracking across 5 days and to a range of clinically relevant tasks (gait, stair navigation, squat, sit-to-stand). These findings support our synchronization approach as a practical route to reliable, multi-day IMU kinematic monitoring in patients with chronic knee pathology.

Because our approach minimizes wearer interaction, it may be well suited to clinical populations with lower technology literacy or adherence, but several challenges remain before deployment in clinical monitoring studies. First, while sensor donning can occur at a clinical visit with the help of researchers, sensor doffing – and the end-of-monitoring physical calibration required for linear resampling – will often need to happen at home before sensors are returned by mail. This may be difficult for patients with reduced hand dexterity and suggests a need for a more user-friendly calibration hardware. Second, our hourly periodic updates occasionally misidentified an offset before self-correcting it at the next checkpoint (Fig. 5). This imperfection could transiently bias knee flexion estimates for up to an hour and may be caused by either imperfect active bout selection or incomplete separation of the shared versus segment-specific motion features. Tuning the frequency of periodic update intervals may help address this weakness. It also remains to be confirmed whether those shared motion features between thigh and shank remain valid for patients with pathological knee motion patterns. Third, our results represent the best case in which no sensor was repositioned during the week, which will not always hold in larger studies. Marking the sensor footprint on the base tape to guide self-reattachment is one practical mitigation strategy, though automatic detection and post-hoc data correction in response to repositioning would be a valuable future addition.

More broadly on the limitations of this study, our synchronization approach was validated only on the AX6. While we expect similar quartz clock behavior in other unconnected low-cost IMUs, we cannot confirm this directly. We did not assess frontal-or transverse-plane knee motion, for which we lacked reliable reference data [24]. The knee flexion estimate is resilient to gyroscope signal drift, but 3-axis Euler angle decompositions would introduce drift-related errors that are not corrected in absence of a magnetometer. Recent work by Phan et al. developed novel strategies to account for these drift errors but still found that knee frontal and transverse plane kinematics produced unacceptably high errors compared against ground-truth biplane fluoroscopy [27]. Finally, our cohort of healthy young volunteers likely adhered to our at-home IMU wearing protocol better than a patient or older population would. Larger studies in such populations should anticipate more non-adherence and practical issues like skin irritation. We nonetheless believe continuous, real-world IMU kinematic data is critical for gaining objective and patient-specific insights into how musculoskeletal diseases and treatments affect movement outcomes in patients during real-world scenarios. Therefore, we encourage continued development of wearable protocols and analytic methods that support multi-day kinematic tracking.

## V. Conclusion

Our study developed a simple and reliable syncing paradigm for low-cost logging IMUs to continuously quantify the knee flexion angle during real-world activities. We showed that our bimodal algorithm reliably syncs two low-cost IMUs to quantify knee flexion for 5 consecutive days, ∼100 uninterrupted hours with comparable accuracy to lab camera-based motion capture. Our 5-day IMU monitoring strategy is a viable solution for clinical studies because it requires minimal wearer interaction and addresses common challenges of patient monitoring with unsupervised software solutions. Our novel syncing algorithms support low-cost wearable IMUs like AX6 to be implemented in at-home experiments on a larger scale. We hope our technical contributions will help expedite the translation of wearable technologies into larger clinical studies, like continuously monitoring multi-day knee kinematics to evaluate how musculoskeletal diseases and treatments affect real-world knee mobility in patients with chronic pathologies like degenerative knee osteoarthritis.

## Data Availability

All data produced in the present study are available upon reasonable request to the authors.

## Appendix

A. Participant Feedback Survey Results

*Total number of participants surveyed:* 15 (all)

*Unfavorable events during monitoring:* mild itchiness = 4; sweat build up inside tape = 1; none = 10

*Other concerns about monitoring:* potential tape loosening or snagging on clothes = 5; sweat or moisture build up inside tape = 3; aesthetic concerns = 1

*Responses specific to thigh IMUs:*

*Overall burden of wearing:* No burden = 7; low burden = 8; medium, high, or very high burden = 0

*Interference with physical activity:* Not at all = 12; rarely = 3; sometimes, often, or all the time = 0

*Interference with sleep:* Not at all = 12; rarely = 3; sometimes, often, or all the time = 0

*Preferred wearing duration in future experiment:* <5 days = 0; 5 days = 3; 7 days = 11; 14 days = 1

*Responses specific to shank IMUs:*

*Overall burden of wearing:* No burden = 7; low burden = 7; medium burden = 1; high or very high burden = 0

*Interference with physical activity:* Not at all = 12; rarely = 3; sometimes, often, or all the time = 0

*Interference with sleep:* Not at all = 10; rarely = 4; sometimes

= 1 (snagging on clothes); often or all the time = 0

*Preferred wearing duration in future experiment:* <5 days = 0; 5 days = 4; 7 days = 10; 14 days = 1

## Acknowledgment

The authors would like to thank Max Zawel for assistance with data collection. We acknowledge the use of artificial intelligence model Claude (Sonnet 5, Anthropic) to assist in editing the text of this manuscript, including its Introduction, Methods, Results, and Discussion. We first developed the full initial draft and then used prompts to reduce redundancy and move non-essential details to Supplemental Material, while maintaining the structure and essential contents of our initial manuscript draft. None of the results, tables, or figures were modified by Claude. We then reviewed, further edited, and verified the manuscript. The authors take full responsibility for all contents in the published version of this work.

