## Supplemental Material for "Physical calibration and signal periodic updates to synchronize IMUs for 5-day continuous knee flexion tracking"

### I. Supplemental Methods S1: Sensor Attachment Protocol

Referenced from main text Section II.C (Sensor attachment).

We attached each AX6 IMU to the skin using a 3-layer medical-grade adhesive system. First, to protect the skin from potential reactions to sensor contact, we placed a 3×2-inch wound-dressing "base tape" (Hypafix, Essity, Stockholm, Sweden) directly on the skin at each attachment site. For participants with more body hair, we offered shaving to reduce discomfort during end-of-experiment tape removal. Second, we applied a double-sided "bonding tape" (Delsys Corp., Natick, MA; originally manufactured for surface electromyography electrodes) to the back of each IMU, then adhered the sensor-tape assembly on top of the protective base layer. We confirmed that the charging port of every sensor faced down and the printed/text side faced the skin, matching orientation across all 4 IMUs – necessary for correctly extracting calibration syncing signals (main text, Section II.E) and sensor-to-segment alignment (main text, Section II.F), and for additional protection from water. We oriented the charging port down to facilitate any water drainage and protect sensor health. Third, we added a 4×3-inch layer of transparent, waterproof, hypoallergenic wound dressing "cover tape" (Opsite Flexifix, Smith & Nephew, Watford, UK) over the entire attachment complex for further protection from water and friction. Most of our study participants reported that the cover tape provided them with peace of mind that their sensor would not fall off. However, the bond between the double-sided bonding tape and the base tape is sufficient to provide secure fixation that lasts for up to 7 days of continuous wear.

We provided replacement cover tape for participants who wanted it mid-week but did not provide replacements for the base or bonding layers; participants were instead instructed to abort the experiment if those layers detached. This occurred for 1 of 15 participants (base tape detachment); this participant was later re-enrolled to complete the full 5-day protocol. For the remaining 14, all sensors remained at their original locations for the full week, with only occasional cover-tape replacement due to wear or sweat buildup, which never affected the underlying sensor position.

### II. Supplemental Methods S2: Physical Calibration Signal Detection and Periodic Update Algorithm – Full Parameterization

Referenced from main text Section II.E (Inter-IMU synchronization: two methods).

#### A. Physical calibration signal detection

We designed and 3D-printed a lightweight rigid holder seating 4 IMUs vertically, following established recommendations for physical inter-sensor calibration [1], [2]. Immediately before the scheduled Monday recording start, we inserted all 4 IMUs into the holder facing upward (opposite their on-body orientation, to produce an automatically detectable signature) and left them static for ≥30 s in this "calibration standby" position, then performed a 3-step rest-swing-rest maneuver: lifting the loaded holder from a flat table, swinging it arbitrarily in the air for ~10 s, then returning it to the table and leaving it static for ≥5 s. We repeated this maneuver on Friday, immediately after sensor removal.

We detected each "calibration standby" interval from two simultaneous threshold criteria in both IMUs: (1) accelerometer signal within -1 ± 0.1g along the local x-axis (i.e., upside-down); and (2) gyroscope local z-axis signal <5°/s (i.e., static). We computed the 5-second rolling sum of samples meeting both criteria and applied a 1-second steady-state threshold to identify entry/exit from calibration standby. The longest such interval was labeled the multi-day wearing period (typically 90–100 h); intervals spanning 6–60 s was treated as candidate calibration maneuvers and were rejected if the two IMUs' gyroscope z-magnitude ranges differed by >200°/s (indicating a possible inadvertent disturbance while inserted in the holder). We selected the longest eligible interval before the wearing period as the Monday calibration signal, and the longest eligible interval after the wearing period as the Friday calibration signal (available for all 15 participants). We used cross-correlation (SciPy "xcorr") on the gyroscope z-axis signal from each calibration interval to identify the inter-IMU time offset.

#### B. Periodic update algorithm

We computed each IMU's resultant acceleration magnitude (2-norm of the 3 axes), subtracted 1g to remove the gravity offset, took the absolute value, and scaled it by each IMU's global recording maximum to reduce the influence of inter-sensor magnitude differences on later cross-correlation. To distinguish active daytime from motionless sleep/rest periods, we computed the absolute value of jerk (the first derivative of resultant acceleration, threshold to zero below 1 g/s to suppress noise), summed thigh and shank jerk, and smoothed the combined signal with a bi-directional 1-hour rolling average over 4 iterations, adapting a method previously described for accelerometer-based sleep detection [3]. We defined the sleep/rest threshold as each participant's global minimum of this smoothed jerk envelope plus 0.025 g/s and retained intervals >1.5 h above this threshold as eligible daytime periods.

Within each eligible active hour in daytime, we defined synchronization checkpoints at full-hour boundaries (pausing overnight). For each checkpoint, we searched forward from the top of the hour for the first 5-minute window meeting two activity criteria – combined-IMU peak acceleration ≥20% of the hourly peak and combined-IMU mean acceleration ≥ the hourly mean – or used the final 5 minutes of the hour if no window met both criteria. Within this window, we applied a 4th-order Butterworth low-pass filter (4 Hz cutoff, negative artifacts reset to 0) to each IMU's scaled resultant acceleration, subtracted each signal's 5-minute mean (demeaning), and cross-correlated the two filtered, demeaned signals to estimate that hour's inter-IMU offset. We discarded implausible offsets using two rejection thresholds: 8.64 s for not-yet-synced data (the reported worst-case AX6 daily drift [1]) or 0.25 s for data already synchronized via linear resampling. If the offset passed this check, we shifted the full-hour unfiltered data of the faster IMU backward (by skipping samples) to match the slower IMU, before advancing to the next hourly checkpoint.

### III. Supplemental Methods S3: Sensor-to-Segment Alignment – Full Axis Derivation

Referenced from main text Section II.F (Knee flexion angle from synchronized thigh and shank IMUs).

We approximated each participant's sagittal knee flexion axis ($\hat{X}$) via principal component analysis (PCA) of each IMU's gyroscope signal during Monday's in-lab squatting trial, during which thigh and shank motion was predominantly sagittal. We used the squatting trial in place of the straight-walking trial recommended in the literature [4] because our short in-lab walkway did not always provide sufficient straight-walking data for the PCA to reliably identify $\hat{X}$. We identified the proximal-distal axis (Z) from the static standing trial, assuming both legs straight and their long axes vertically aligned with the static accelerometer signal (gravity).

To correctly locate the squat and standing-pose windows within the continuous, unlabeled IMU recording using the motion-capture (ground-truth) clock, we corrected for the IMU-to-lab clock offset – distinct from the inter-IMU synchronization described in the main text – by cross-correlating the generally sagittal gyroscope local-z signal against segment sagittal velocity computed from motion-capture data during the squat trial, and by removing any explicitly known offset between the two computers used for motion capture and IMU setup.

We then derived the antero-posterior axis (Y) as the cross product of the flexion axis ($\hat{X}$) and gravity (Z) and refined the final sagittal axis (X) as the cross product of Y and Z, yielding a full orthogonal anatomical frame into which we virtually rotated each IMU's accelerometer and gyroscope data.

### V. Supplemental Table S1: START AND END DEFINITIONS OF MOVEMENT REPETITION DURING IN-LAB MOTION CAPTURE TRIALS

Referenced from main text Section II.C (In-lab motion capture).

| Trial Name | Movement Starts | Movement Stops |
|---|---|---|
| Straight walking | Heel strike | Subsequent heel strike (i.e., gait cycle) |
| Arbitrary walking | A standing position | Reaching 30 seconds of arbitrary motion |
| Squat | Center of mass starts descending | Center of mass stops ascending |
| Step up | Side of interest foot lifts off lower step | Contralateral foot touches upper step |
| Step down | Contralateral foot lifts off upper step | Side of interest foot touches lower step |
| Sit-to-stand | Center of mass starts ascending | Center of mass stops descending |

### VI. Supplemental Figure S1

Knee flexion angle tracking results when using linear resampling only are shown below in Fig. S1.

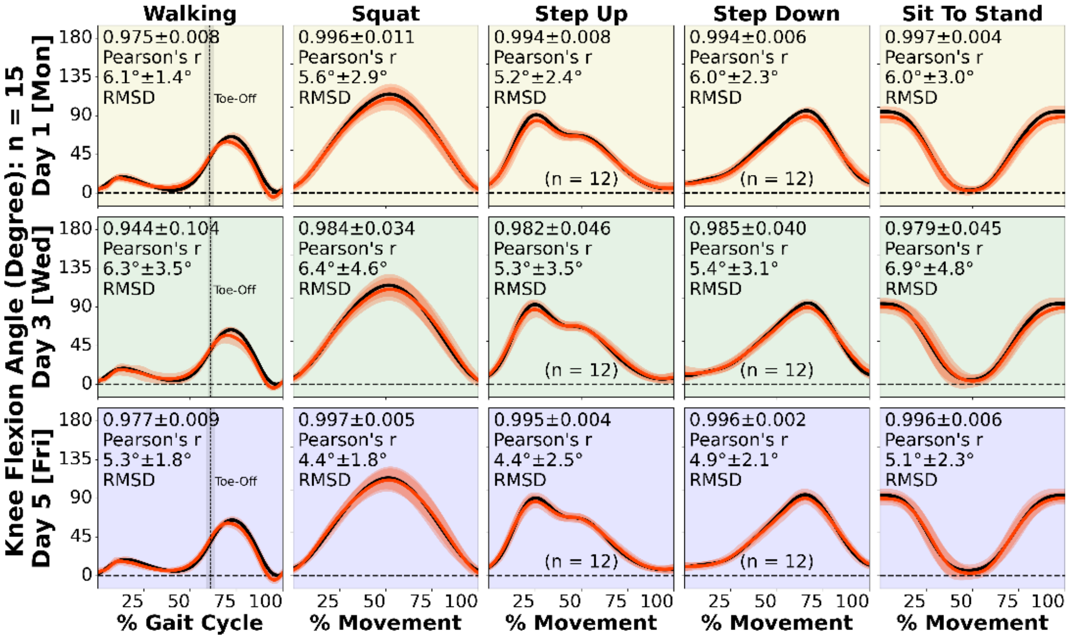


**Fig. S1.** Group-wise mean (line) ± 1 standard deviation (shade) on IMU-estimated knee flexion (red) versus camera-based motion capture (black) on Monday (top), Wednesday (center), and Friday (bottom). Data shown was synced using calibration-based linear resampling ONLY. Although these group-level results were similar compared to periodic updates-based methods (Fig. S2; Fig. 9 in main text), we observed multiple individual outlier cases where mid-week tracking errors were substantially worse and unacceptable (Fig. 8a in main text).

### VII. Supplemental Figure S2

Knee flexion angle tracking results when using periodic updates only are shown below in Fig. S2.

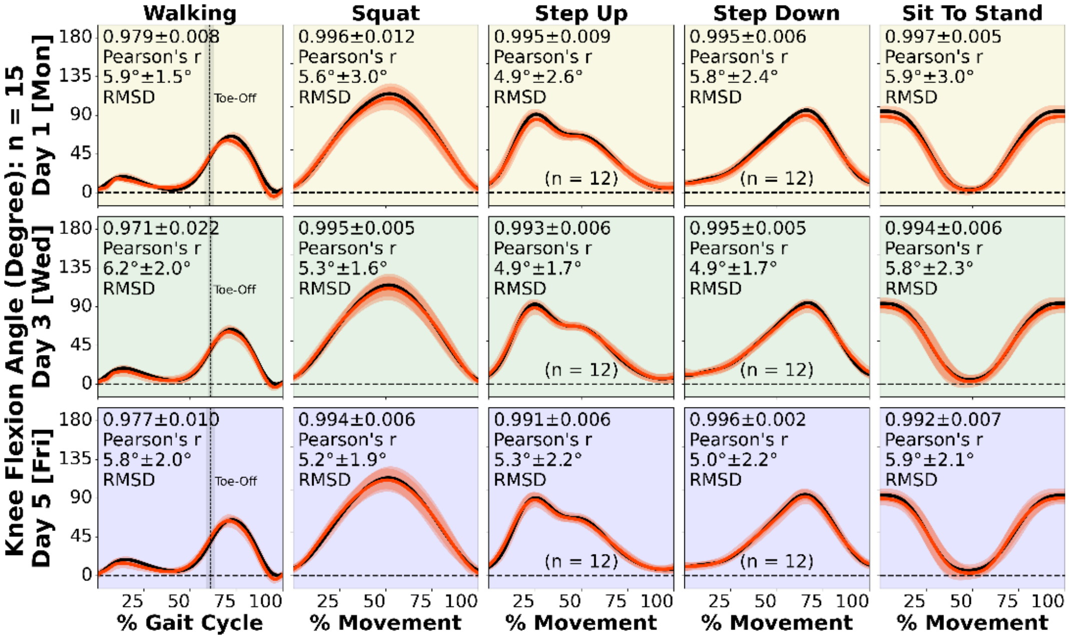


**Fig. S2.** Group-wise mean (line) ± 1 standard deviation (shade) on IMU-estimated knee flexion (red) versus camera-based motion capture (black) on Monday (top), Wednesday (center), and Friday (bottom). Data shown was synced using signal-based periodic updates ONLY. These results were largely equivalent to using periodic updates alone for syncing (Fig. 9 in main text), while advantageous to using linear resampling alone at the individual level, as outlier cases of substantial mid-week tracking errors were no longer observed (Fig. 8b in main text).
